# Keen relationship between COVID_19 and food supply suggest some animal origin foods as excellent vehicle of SARS-Cov-2

**DOI:** 10.1101/2020.06.16.20132464

**Authors:** Nazli Saeedi, Seyed Abbas Rafat

**Affiliations:** Research Center for Pharmaceutical Nanotechnology, Biomedicine Institute, Tabriz University of Medical Sciences, Tabriz, Iran; Department of Animal Science, Faculty of Agriculture, University of Tabriz, Tabriz, Iran

## Abstract

In recent months, nearly all countries tried to decrease human-to-human contact as the principal mode of transmission of SARS-CoV-2. However, other modes of transmission also need to be clarified in more depth, especially, the foodborne transmission. We assessed the effect of animal origin foods consumption on the pandemic of COVID-19. For this purpose, we studied the relationship among 20 food supply as independent variables, and the parameter of Total Cases as dependent variable. Here we show a relationship between a group of animal origin foods and total cases. Regression, Bayes, and Lasso results showed that eggs and fresh water fish have positive coefficient. So, among the transmission ways of COVID_19, the role of foodborne transmission should be more significant than previously thought. The possibility of animal origin foodborne transmission should be taken into more consideration. The perspective is to expand the surveillance of SARS-Cov-2 during the food production chain. In conclusion, the results of the present study indicate that one important vehicle for SARS-Cov-2 may be some of animal origin foods. It is recommended that virologists examine the possibility of freshwater fish and chicken‘s eggs being as excellent vehicles/preservatives for SARS-Cov-2.

## Introduction

At the time of writing this manuscript, the number of total cases of COVID-19 past 28 mark. During the pandemic, most countries around the world tried to minimize human-to-human contact and travels(Merler et al., 2020). However, the prevalence of COVID-19 continued. Despite efforts of many countries to reduce the transmission by person-to-person way the rapid outbreak shows the necessity of research on other possible transmission routes. During early pandemics, in Italy and the USA, for example, the chances of being surprised by pandemics were low, but both countries faced epidemic dramatically. Among the various routes of transmission, FBT is most likely. We suppose the role of COVID-19’s transmission by other ways should be more significant than previously thought. Transmission of MERS, a similar pathogenic virus to SARS-Cov-2, through food has strengthened our hypothesis.

Furthermore, some researchers cited the possibility of FBT for SARS-Cov-2(Hi, 2020; Kingsbury et al., 2020; Newell et al., 2010; Panel and Biohaz, 2011; Todd et al., 2020). Also, FAO/WHO experts mentioned the data gap in the relationship between viruses and food borne disease (FAO/WHO, 2008). The motivation for the present study is to find the effects of human food consumptions (independent variables) on COVID_19 (As dependent variable).

## Material and Methods

Table 1 shows descriptive statistics of studied variables. We assessed the possible relationship between the consumption of main foods and total cases od covid_19. Accordingly, we studied total cases as a dependent variable and 20 food supply data as independent variables. The primary purpose of the study was to find effect of anima origin food on total cases, however, we considered six plant origin foods for comparison reasons. The statistical methods included correlation, stepwise and LASSO regression, and Bayes analysis. Food supply (kg/capita/yr.) was quantity in each country based on the latest available data of FAO. Adjusted R-square of fitted models are presented in Table 2, respectively. Detail material and methods are included in supplementary materials.

**Table 1.**
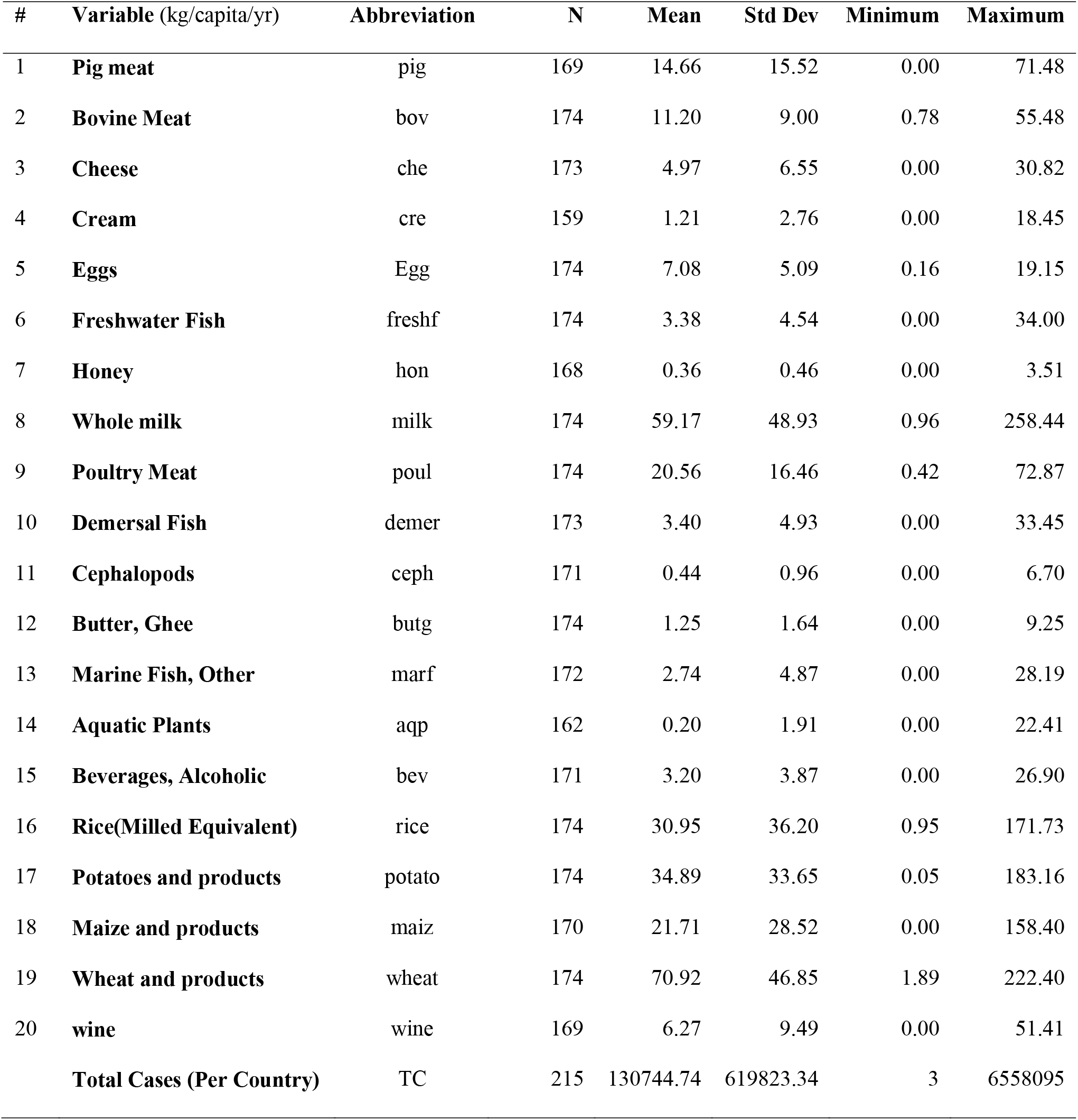
Descriptive statistics of studied foods (n=20) besides total cases.

**Table 2.** R-Square, Adjusted R-square, AIC and number of data in the fitted models for food consumption.

|  | **Stepwise** | **LASSO** | **BAYES** |
| --- | --- | --- | --- |
| **R-Square** | 0.33 | 0.47 | - |
| **Adj-R Square** | 0.31 | 0.42 | - |
| **AIC** | - | 313 | 562 |
| **n** | 131 | 131 | 130 |

We estimated the relationship between 20 food supply and Total Cases of covid19. The methods include Stepwise Regression with Proc Reg, LASSO regression with GLMSELECT Proc, and Bayes analysis with Proc Genmod (SAS). Food supply was quantity (kg/capita/yr.). Fourteen foods of animal origin and six foods of plant origin were selected. The primary purpose of the study was to find AOF relationship with total cases, but some plant origin foods were chosen for comparison reason.

We started our analyses first time on March 2020. Then we repeated it several times with updated COVID_19 data on April, May, June, and September. Finally we presented the latest results.

## Data

We downloaded the data the following websites: COVID_19: https://www.worldometers.info/coronavirus/ 10 September 2020 Crops Primary Equivalent: http://www.fao.org/faostat/en/#data/CC Food Supply - Livestock and Fish: http://www.fao.org/faostat/en/#data/CL

We used base-e logs, also known as natural logs for the transformation of total cases and independent variables. In all analyses, the accuracy of the model with non-transformed data was very low. So, all variables has been transformed to normalize the distributions. With transformation of data, the accuracy of the model increased to an acceptable level. In comparison of models, Lasso’s results are more reliable than stepwise regression and Bayes, because it’s AIC was smaller and also it had higher accuracy as R-square (Table 2). Visual examination of the trace plot displays proper mixing in Bayesian analysis.

We provide the SAS code for an application of the used statistical methods (Rafat, 2020). We note that this work was posted as preprint on medrxiv (Rafat, seyed abbas, 2020).

## Results and Discussion

Stepwise Regression, Bayes, and Lasso’s results confirmed that eggs and fresh water fish have high positive relevance with total cases. With our results in Table 3, implying the need for tracing of SARS-COV-2 in main AOF. Bayes results, e.g., show there is a 0.99 probability of a positive correlation between eggs and total cases, adjusted for the other covariates. The number of AOF and their role in our results varies depending on the type of statistical method used, but all results are consistent. The results of all three statistical methods confirm each other in the case of eggs. Negative high correlation between marine fish and total cases is curious observation needs more study.

**Table 3.**
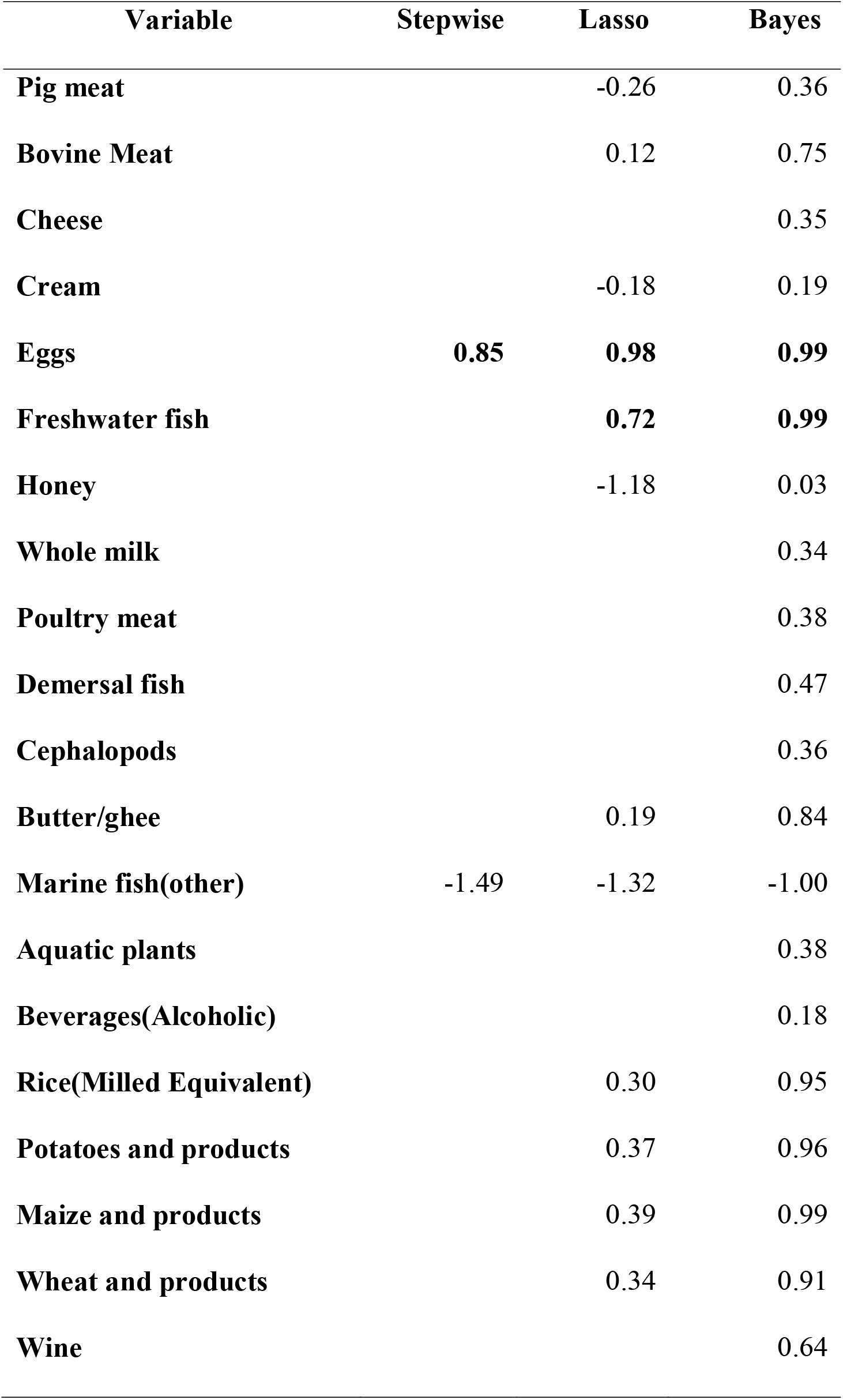
The results of multivariate analyses. All variables are logarithmic transformed data.

The effect of a group of AOF on total cases was significant in our analysis. So, we suppose a noticeable effect of AOF as FBT/vehicle. At present, we are not able to interpret all significant effects and their coefficients. Interpretation of the factors could be the subject of future researches. Potential transmission ways of COVID-19 based on the results of our analysis and bibliography is summarized as Fig 1. Lake and Kingsbury (2020) provided a review about potential for FBT of COVID-19, but they could not guarantee the application of the data found on other coronaviruses to SARS-CoV-2(Kingsbury et al., 2020). The persistence of viruses by refrigeration temperature(FAO/WHO, 2008) might be an explanation of the relationship between COVID-19 and AOF. The viral persistence of coronavirus in soft cheese (Panon et al., 1988) and raw milk(Hamdi, 2013) has been reported. We don’t know what’s going on from “Farm to Fork” about AOF. For example, SARS-CoV-2 may be more resistant to heat when cooking, as it is observed for poliovirus in beef(Pirtle, 1991). It is recommended not to consume raw or undercooked animal products(Moy, 2020). Kitchen utensils can transmit the virus among foods(Sarah C. P. Williams, n.d.), accordingly, AOF may be a suitable vehicle, and the virus inside them will survive better and longer.

**Fig. 1.**
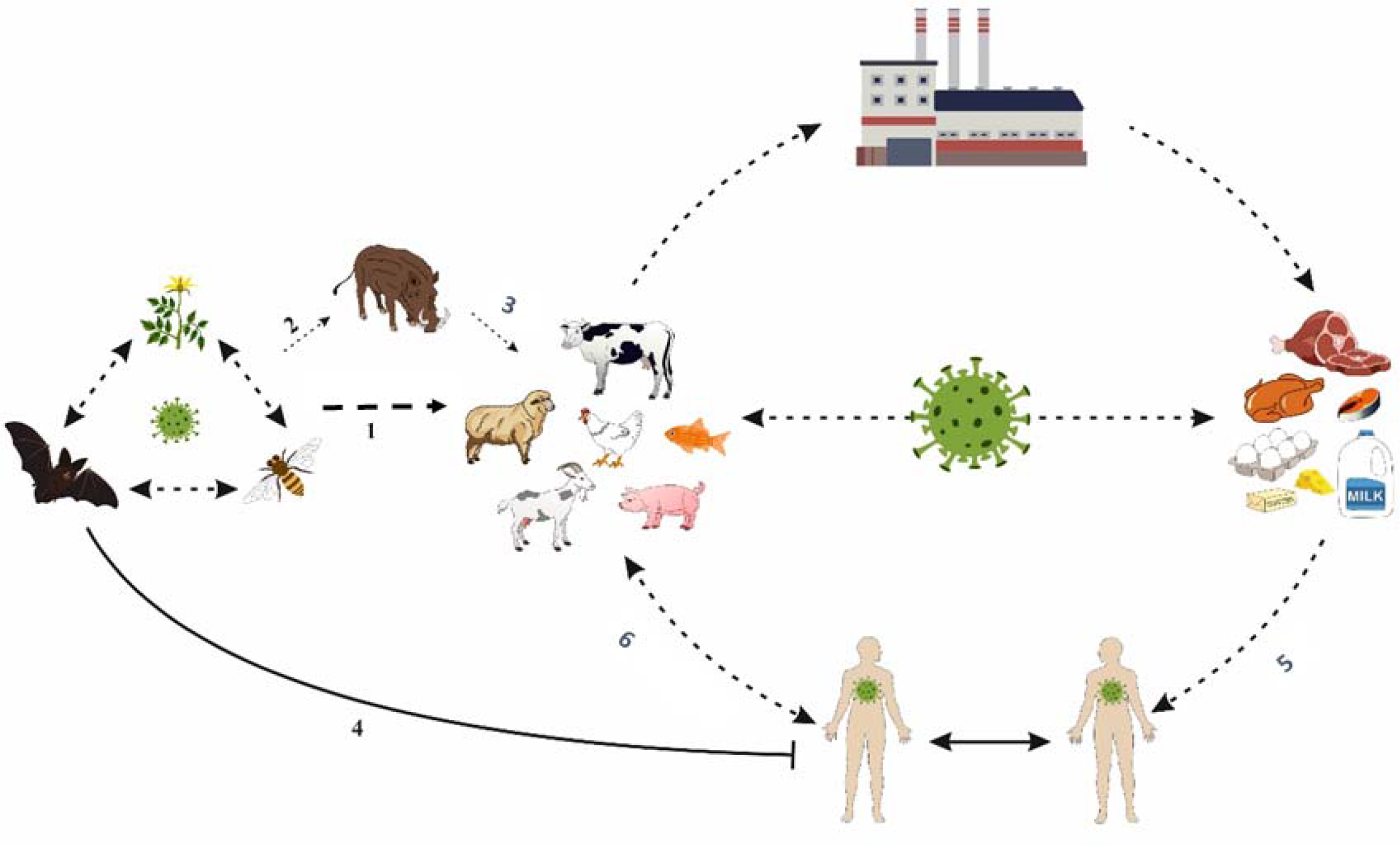
Potential transmission ways of COVID-19 based on the results of our analysis and bibliography. It is necessary to study the exchange of human pathogen viruses in wildlife and livestock. Where and how suspicious foods become contaminated must be investigated. The potential spillover of pathogenic viruses from arthropods hosts to humans should be investigated(Tao et al., 2020). The cross-species transmission is reported(Miller et al., 2017). There may be a type of contact among flowers, bees, and bats. Infected humans could have transmitted MERS to the camels (Kai Kupferschmidt, 2013), similar situation is imaginable for SARS-Cov-2. SARS-CoV-2 and MERS-Co are among the most notable examples of spillover into humans(Guan et al., 2003). References for arrows: 1 (Bourgarel et al., 2019; Cavanagh and Cavanagh, 2007; Maori et al., 2019; Mcmahon et al., 2018; Moy, 2020; Rizzo et al., 2017) 2: (Miller et al., 2017; Moy, 2020) 3: (Miller et al., 2017; Papineau et al., 2019) 4:(Geisbert et al., 2014; Tao et al., 2020) 5: (Gibbins et al., 2020; Newell et al., 2010; Panon et al., 1988) 6: (Gibbins et al., 2020; Hi, 2020; Katy Askew, 2020)

Furthermore, the methods of diagnosing the virus in food are not complete, and there are many limitations(Bosch et al., 2018). In food science technology, the primary attention paid to bivalve moullouses. Nowadays, it is necessary to expand the detection of viruses to all ANIMAL ORIGIN FOOD.

The high-fat tissue in the body has effect on the pathogenesis of COVID-19(Maffetone and Laursen, 2020). Although the probability of transmitting the virus through wine is zero, we deliberately kept it in the initial model of our analysis. Results showed wine is eliminated among significant variables of the suited model (Table 3). So we concluded that just the energy content of food should not keep a variable in the fitted model. So, in addition to energy content, there must be something else in food-related COVID-19. We concentrate on the transmission of COVID-19 by AOF, rather than the immunity response.

The paper of (Katy Askew, 2020) guided us to construct our hypothesis about the essential relationship between COVID-19 and AOF include.

WHO’s recommendations state that COVID_19 have not FBT, and suitable food safety practices could prevent the probability of FBT. However, the realization of proper food safety practices for all AOF is questionable. It is reported that acquisition of SARS coronavirus by consumption of contaminated food is possible (Yepiz 2013). Furthurmore, oral infection of SARS-Cov-2 could not be excluded among transmission ways (FAO/WHO 2008). Promkuntod 2006 recommends taking precautions to prevent the spread of the avian flu by consumption of quail eggs(PROMKUNTOD et al., 2006). One explanation for our observation is the trapping and accumulation of sars-cov-2 by AOF. Thereafter, along food processing or preparation, AOF conserve SARS-Cov-2 infectively.

## Conclusion

Despite the awareness and readiness of many countries for COVID-19 during early 2020, why did it spread so quickly? It seems FBT has received less attention relative to other routes. The perspective is to expand the surveillance of SARS-Cov-2 during the food production chain. It is recommended that virologists examine the possibility of freshwater fish and chicken‘s eggs being as an excellent vehicle/preservator for SARS-Cov-2. Some animal origin food may have been the first cause of transmission of the SARS-Cov-2 from reservoirs populations to humans.

## Data Availability

The code including data used for our analyses is publicly available https://www.researchgate.net/publication/344270580_data_of_Keen_relationship_between_COVID_19_and_food_supply_suggest_some_animal_origin_foods_as_excellent_vehicle_of_SARS-Cov-2_httpswwwmedrxivorgcontent1011012020061620132464v4

https://www.worldometers.info/coronavirus/

http://www.fao.org/faostat/en/#data/QA

http://www.fao.org/faostat/en/#data/CC

http://www.fao.org/faostat/en/#data/CL

## Author contributions

Conceptualization and Visualisation: N. S. K., Methodology: S. A. R. Writing: Both authors. The authors have declared no competing interest. Data and materials availability: All data are available in the manuscript or the supplementary materials.

